# Individual and system causes of moral distress experienced by public health practitioners in Canada

**DOI:** 10.64898/2026.06.02.26354688

**Authors:** Jenna Bennett, Smita Pakhale, Nicola Desmond

**Affiliations:** The Ottawa Hospital Research Institute, Ottawa, Ontario, Canada; Department of Global Health and Development, London School of Hygiene and Tropical Medicine, Tavistock Place, London, United Kingdom

## Abstract

**Aims:** Moral distress has been studied across many health arenas; however, public health has often been overlooked. Canada is facing a healthcare crisis with a significant number of staff leaving the healthcare field. This study explores the experiences of moral distress in public healthcare practitioners across Canada. Better understanding these experiences can provide insights into how to support staff and prevent attrition in public health.

**Methods:** This was a cross-sectional qualitative study. Fifteen in-depth interviews were conducted between May and July 2023, through remote and in person methods. Participants were from nursing, social work, medicine, and dietetics, all working in public health across Canada. Iterative thematic analysis was used. Emergent themes were compared within and across data sets and by participant age and years of experience.

**Results/Findings:** Experiences that contributed to moral distress included systemic powerlessness, political and ideological overreach, unethical work environments and undervalued expertise. Years of experience and diversity in gender and ethnicity impacted how practitioners navigated moral distress. Experiences where practitioners felt actions went against their values increased during the pandemic, contributing to moral injury.

**Conclusions:** This study situates the unique position of public health within the health system and explores experiences of moral distress both during and outside the COVID-19 pandemic. While the pandemic brought the concept of moral distress to the forefront of many people’s minds, these experiences existed prior. Addressing the underlying causes will contribute to establishing approaches to support public health practitioners suffering from moral distress and injury.

## Introduction

The COVID-19 pandemic exposed widespread moral distress among health practitioners and significant workforce shortages in high-income countries like Canada. In the third quarter of 2024, Canada reported 78,600 unfilled healthcare positions [1]. A Canadian study reported a 78.7% burnout rate among public health workers, among the highest globally, with burnout significantly increasing the likelihood of early retirement or departure [2]. Since moral distress contributes to burnout and attrition, understanding its causes is vital for workforce sustainability [3]. While moral distress has been extensively studied in clinical settings such as palliative and critical care [4,5], research in public health remained limited before the pandemic when it was recognised as a significant problem, with potential to cause harm to individual professionals, the workforce and ultimately the wider population [6–8].

‘Powerlessness to influence discriminatory systems’ is a key driver of moral distress in public health practice [9]. Public health identifies moral aims of advocacy and social justice, focusing on both upstream and downstream work [10,11]. During the pandemic practitioners witnessed systemic failures while tasked with both front-line care and informing systems level decisions such as vaccine mandates and isolation policies. These measures disproportionately burdened marginalized populations [7], forcing practitioners to navigate complex individual ethical dilemmas within systems approaches [10]. Disconnects between system wide interventions to minimize harm to individuals often lead to violent backlash against public health officials [12].

Although several ethical frameworks aim to reconcile the individual-centred focus of bioethics with the population-level concerns of public health ethics [10], such as ‘Looking ahead: Addressing Ethical Challenges in Public Health Practice” [13,14] they often lack practical guidance and fail to reflect everyday moral experiences, leading to confusion, apathy or even unethical behaviour [10,14–16].

Moral distress was first introduced into healthcare in 1984 by Andrew Jameton as “the negative experience when one knows the right thing to do, but institutional constraints make it nearly impossible to pursue the right course of action” [17]. The concept has since evolved to include moral residue, ‘lingering feelings after a morally problematic situation has passed” [18,19] and moral injury “perpetrating, failing to prevent, or bearing witness to acts that transgress deeply held moral beliefs and expectations” [18,20–22]. Moral outrage; “anger provoked by a real or perceived violation of an ethical standard such as fairness, respect, or beneficence” [23] has been identified as an outcome of moral distress and injury. The concept of constraints is consistently present in definitions [19,24–27], with internal constraints shaped by experiences, beliefs and values, and external constraints arising from systems and institutions [25]. In public health, external constraints are compounded by political structures [25]. In Canada, many public health practitioners work in government-funded agencies, limiting their ability to challenge policies and structures [11]. Public health’s dual focus on upstream and downstream interventions creates overlap with systems beyond healthcare, creating a unique space to affect change, while also creating morally distressing experiences distinct to public health [28].

Although recent research has begun addressing moral distress in public health, it remains largely survey-based and pandemic-focused. This paper presents in-depth experiences of moral distress amongst public health practitioners and situates this within their unique position in promoting health at both the individual and systemic level creating experiences of moral distress that are unique to this space.

## Methods

This is a cross-sectional, qualitative study exploring the lived experiences of moral distress and ethical dilemmas among public health practitioners in Canada.

Recruitment material [posters and handouts] was shared through Twitter, Facebook, LinkedIn, and physically posted in locations where public healthcare practitioners were known to work in Ottawa, Canada. Organizations connected to Canadian public health professionals disseminated recruitment material including during a June 2023 Canadian Public Health Association webinar on moral injury [https://canvax.ca/canvax-webinar-series#w13]. While participants self-identified we purposively recruited ethnically diverse staff through targeted approaches and snowball sampling. Potential participants were screened for licensure as health practitioners. One interested participant, a health promotion officer, was excluded.

Fifteen in-depth interviews were conducted through May to July 2023 by J.B, a female Master’s in Public Health student and Canadian registered nurse with 15 years of public health experience. Fourteen interviews were remote; one was in-person at the participant’s workplace. Interviews lasted 60 to 90 minutes, were audio recorded, transcribed and accompanied by field notes. Basic socio-demographic details and participant understandings of moral distress were collected before introducing the formal definition, followed by an open-ended question on the experiences participants wished to discuss. A discussion guide based on moral distress mapping covering emotions, sources, constraints, responsibilities, and actions was used [Data in S1 Discussion Guide] [29].

Iterative thematic analysis was conducted, and data collection ended following saturation. Interviews were coded manually with content driven descriptive codes with definitions, details of use and code demonstrations from the interview text. Interviews were reviewed to confirm and adjust coding and to identify themes. Inter-code and theme relationships within and across transcripts were explored. Data sets were reviewed based on age and years of experience. All coding and themes were discussed between J.B. and N.D. Where possible, data contrasts pandemic and non-pandemic experiences of moral distress.

### Ethical considerations

Ethics approval was received from the London School of Hygiene and Tropical Medicine, reference # 28455; and The Ottawa Hospital Research Institute – Ottawa Health Science Network Research Ethics Board, protocol # 20230120-01H.

### Consent to participate

We obtained verbal informed consent for remote interviews and written consent for the in-person interview following review and discussion of study information. Verbal consent received ethical approval and was documented in transcripts.

## Results

Participant characteristics and years of experience are presented in Table 1. Participants included licensed healthcare practitioners from nursing, medicine, dietetics, and social work, all self-reporting practice in public health in Canada. Those identifying as public health nurses accounted for 86% of participants, reflecting their majority within the healthcare sector [30]. The sample included those practicing in 5 Canadian provinces, in both rural and urban settings.

**Table 1.** Participant Characteristics and Years of Experience.

| **Participant Characteristics** | **Frequency [%]** |
| --- | --- |
| **Gender Identity** |  |
| Female | 13 [86.6%] |
| Other | 2 [13.3%] |
| **Age** |  |
| 25-34 | 3 [20%] |
| 35-44 | 6 [40%] |
| 45 and over | 6 [40%] |
| **Race** |  |
| White | 13 [86.6%] |
| Other | 2 [13.3%] |
| **Years of Experience as a Healthcare Practitioner** |  |
| 0-10 | 3 [20%] |
| 11-20 | 7 [46.6%] |
| 21 and over | 5[33.3%] |
| **Years of Experience in Public Health** |  |
| 0-5 | 5 [33.3%] |
| 6-15 | 5 [33.3%] |
| 16 and over | 5[33.3%] |
| **Higher education** |  |
| Undergraduate degree only | 4 [26.6%] |
| Masters or Graduate degree [completed or completing] | 11 [73.3%] |
| Multiple degrees/diplomas [including master's or graduate degree] | 5 [33.3%] |

Moral distress was associated with systemic powerlessness, political and ideological overreach, unethical work environments and undervalued expertise. Moral injury emerged through discussions of moral distress. Years of public health experience helped manage moral outrage by channelling anger into advocacy. Inequity as a systemic cause of moral distress was explored through those identifying as a minority due to gender identity or race. Results are presented by theme, with participant quotes.

### Systemic powerlessness

Systemic powerlessness was foundational in many experiences of moral distress, felt in decision-making, policy direction, communication and scope of work. Leadership styles were described as dictatorships, ignoring staff voices. Promotions helped improve moral distress, while leaving leadership roles exacerbated it.

> *And so, you know, as a nursing project officer [a promotion], people listen to me a lot more than they did when I was a public health nurse, even though I’m the same person with the same level of knowledge and the same level of experience and the same level of expertise. And so, I found that frustrating. I’m glad they listen to me now, but I shouldn’t have had to get a promotion for people to listen to what I had to do or what I have to say, you know. [P001, Nurse, 15 years in public health, 17 years in healthcare]*.
>
> *I think for me another source of moral distress is really as someone who was in a leadership position when I could influence decisions, no longer being in that position and no longer being able to influence decisions about where our programs are targeting. I do feel distressed because I often wonder who is at the table now. [P008, Nurse Practitioner, 11 years in public health, 22 years in healthcare]*.

Distress deepened when leadership limited or removed programming staff felt was needed.

> *Supervisors within our institution who don’t even feel that we should be providing direct clinical services because we’re public health. So, in my stance like that, that upsets me because I know that our services are needed. So, when I hear someone in a leadership position or someone who makes decisions kind of wanting to pull back our healthcare services, umm, you know, I definitely feel very stressed myself, and wondering like how can I kind of advocate for this. [P008 Nurse Practitioner, 11 years in public health, 22 years in healthcare]*.

HIV programming was given as an example where program funding was tied to basic HIV education, excluding risk factors like oppression and intimate partner violence.

> *[…] they had a very strict way of financing services and really narrow vision of what type of work had to be done. It was mostly centered around knowledge. The knowledge growing, but they’re in a very narrow way and narrow view of it. It was not structurally informed, it was not trauma informed. It was really like basic HIV knowledge you have to know and that would fix everything else, and that was very confronting with all the minority stress theory that’s been growing in the recent years to move beyond just the HIV paradigm and to really view health in a global manner with social determinants, with oppressions, and beginning to see that to prevent HIV you had to care about mental health, you had to care about intimate partner violence […] if you put on services and activities and the way you make your questionnaires or things that don’t produce the results that are expected, those very outdated type of expectations, then you’re not doing your job and we’re not funding you and there’s always the menace of cutting funding. [P004, Social Worker, 5 years in public health and healthcare]*.

Powerlessness also extended beyond health systems or funding – for instance, in Catholic schools, sexual health education had to be limited to avoid being barred from returning. Moral distress was experienced as practitioners couldn’t provide the care they felt was needed and knew how to perform.

> *And the question that gets asked in the question box, the anonymous question box, is ‘what makes somebody a girl or a boy and can somebody be in between?’. And I’m in a Catholic school and they’ve just started inviting us in and they’ve warned us that we’re not to talk about transgender issues. We’re not to talk about abortion. We’re not to talk about masturbation. We’re not to talk about same sex marriage. All of these things. [P010, Nurse, 15 years in public health and healthcare]*.

During the pandemic, powerlessness intensified due to poor communication and lack of information, hindering care and contributing to moral distress. Practitioners faced complex situations such as vaccine rationing with limited and outdated information, contributing to what they described as an ‘erosion of public trust’ in their capacity to provide care.

> *There was lots of times in public health […] where people would come to get vaccinated and they would be the ones to tell you that the policy had changed overnight because they read it in CBC News. We didn’t know it on the front line and that continually made us feel and made us look stupid. Made us feel like why were we the last ones to know? We should have been the first. It eroded our perception of professionalism […] it feels like it eroded public trust. [P011, Nurse, 3 years in public health, 12 years in healthcare]*.

Participants used statements such as “we were sacrificed” or “we were thrown to the wolves”.

> *I wish […] I had more information, or I had better support. Just in general, they kind of threw us to the wolves, right? [P002, Nurse, 2.5 years in public health and healthcare]*.

Information limitations contributing to feelings of powerlessness were seen as specific to public health, as other healthcare fields strive to communicate evidence. Political overreach further fuelled powerlessness by controlling information and limiting care delivery.

> *Even though it is evidence based does not mean that public health can incorporate it. When you’re in acute care, you want evidence based everything right, best practice. Well, best practice in a political realm is not necessarily going to fly. You need to have the groundswell. [P003, Dietician, 17 years in public health, 28 years in healthcare]*.

### Political and ideological overreach

In public health, politics influences budgets, resource allocation, and programming, contributing to practitioners’ moral distress. In Canada, healthcare decisions are made provincially, with varying municipal and federal involvement. Political ideologies shape health policies and participants described suppression, intimidation, and job threats when programming conflicted with political agendas.

Provincial politics particularly affected participants working on poverty reduction in conservative political contexts leading to frustrations which in turn led to moral distress. For example, a report recommending a minimum wage increase to improve food affordability was suppressed. Its authors faced reprimands and professional threats, leading to anger and resentment that the information was withheld.

> *[…] I think on page 17 we had a list of, you know, what can a community do? What can a municipal government do? What can a provincial and federal government do? And one of them was increased minimum wage. And it caused, we had our hands slapped, we were told we couldn’t do this anymore […] I think the emotion that came up, when it was slapping my hand, was determination. Umm. Anger. Resentment. Umm. And I think […] that sense that people need to know this info, and I’ve got it, and we worked hard on it, but I don’t have control of what happens. [P003, Dietician, 17 years in public health, 28 years in healthcare]*.

Some participants vocalized that in public health, health had less of an influence on decisions than politics.

> *So people often make ties between legality and morality, even though they’re two separate things. And so, because of that inherent point between legality and morality that affects politics, that affects how people vote and then that affects policy and practice, oftentimes in healthcare. And when you work in a municipal government, it does, you know, at the end of the day, like the city is elected officials. [P009, Nurse, 17 years in public health, 24 years in healthcare]*.

Budget cuts linked to political decisions deepened moral distress, leading to staff disillusionment and attrition, resulting in a loss of expertise witnessed over years.

> *Sure, the cuts have hurt, but the cuts remove brains from public health. They don’t remove material, they don’t remove MRI, they remove expertise, they lead to people retiring earlier to a wealth of experience and expertise and wisdom and how to navigate those complex pathways in public health being removed quite suddenly because there’s a vicious circle. If there’s a big cut, the morale is low. People will accelerate their departure, for whatever reason. [P007, Physician, 24 years in public health]*.

During the pandemic, political discourse devalued public health knowledge. Situations were discussed where local chief medical officers would make politically reassuring but scientifically unsound statements.

> *But to see how on a daily basis I could witness how the expertise was not being valued, not being recognized, and sometimes being actively pushed away because it was not convenient politically, has been another, umm… test of reality, I would say. [P007, Physician, 24 years in public health]*.

This pattern continued as the individual occupying this position changed, pointing to political pressure rather than individual failings. Intimidation was witnessed, not just by individuals, but by whole political communities.

> *When intimidation is one person to work towards one person, there are some tools. You can witness, you can reflect, you can support the person who is being intimidated, you can call the intimidation. When it’s one person against a pack, which is the case when there are political muzzling, silencing mechanisms at play. There is no easy way out. [P007, Physician, 24 years in public health]*.

Witnessing significant pandemic spending at the expense of programs supporting vulnerable populations resulted in moral distress. For example, despite Canada’s ongoing opioid crisis [31], addiction programs struggled with funding.

> *I think resource allocation was a big one. Yeah, I think, like epi data shows fatalities, right? So, it’s like we always use epi data to justify where our money goes or where resources are allocated. But you wonder, like you know, at this point in the game, like the COVID money or the COVID fatalities are very low. And there was just an article published, I think last week or a few weeks ago about how in British Columbia overdoses were the number one cause of death between the ages of […] 20 to 59 or something and it was more so than suicide and accidents and all these other things combined. So, it’s like, you’re kind of just like, what? What do they have to do besides die to justify this mass influx of help? [P009, Nurse, 17 years in public health, 24 years in healthcare]*.

Those working in rural communities witnessed racially driven resource failures, worsening inequities.

> *There were so many challenges there and like such a lack of resources that I was constantly surprised like this is [my province] that is connected by roads and this is the treatment and this is how robust we can have any kind of healthcare for this population? [P012, Nurse, 7 years in public health, 11 years in healthcare]*.

While resource constraints were expected, politically driven decisions that perpetuated systemic discrimination were especially morally distressing. Advocacy efforts were often met with threats, leaving participants feeling their practice had become unethical.

### Unethical work environments and undervalued expertise

Inequitable resource allocation, organizational focus on rhetoric over action, and budget cuts targeting health promotion, prevention, and vulnerable community programs contributed to feelings of undervalued expertise and unethical work environments.

Participants described being placed in settings they viewed as unethical, as organizations prioritized politically safe programs over meaningful equity work.

> *And so, I find it really interesting that we say as an organization and we pat ourselves on the back for valuing diversity, equity and inclusion, yet refusing when it’s not convenient for us to do something. It’s only, we only value those things when it’s convenient for us to value them and easy for us. [P001, Nurse, 15 years in public health, 17 years in healthcare]*.

Budget cuts exacerbated feelings of undervaluation and reduced capacity for prevention, fuelling competition for limited resources instead of encouraging collaboration.

> *And it ends up, you know […] it can fracture some of these relationships or make them awkward when really all of us, I’m pretty sure, have the same goal. But it doesn’t seem like we’re getting there together. We’re fighting each other as we’re trying to, like, win together. It’s really odd. [P012, Nurse, 7 years in public health, 11 years in healthcare]*.

While practitioners had seen budget cuts for years, their impact was more apparent during the pandemic, constraining efforts in prevention.

> *Just in general a disillusion and a realistic observation that our societies, our Canadian society, are not dealing with prevention in a significant way. It’s not the main focus. We have relatively little investments or minimal investments in public health compared to many other realms of health funding. So that would be my first area of major disillusion and how it was lived, very practically, during the pandemic. The lack of funding, the cuts that we have had. In [my province], the regional public health units were cut one third of their budget in 2015, one third. That was immense and it had profound implications on our ability to respond to this pandemic or just to function as an organization since 2015. [P007, Physician, 24 years in public health]*.

As pandemic priorities took precedence, feelings of being undervalued deepened. For example, amid practitioners’ recognition of rising cases of shaken baby syndrome and infant injuries, some programs supporting vulnerable families were drastically reduced, with staffing cut from 17 to 2.

While perceptions of unethical work environments and undervaluation of expertise existed previously, they were exacerbated during the pandemic, pushing moral distress into injury.

### Moral injury

When discussing experiences, many participants described experiences of moral injury. These included vaccinating hesitant individuals, experiencing violence from those they aimed to help, and working in harm reduction while witnessing repeated self-harm without systemic solutions.

Work during the pandemic in unsupportive environments while managing complex ethical dilemmas was repeatedly discussed. For example, while there was support for vaccine mandates, establishing a balance between promoting the collective good without having time to establish true individual consent placed practitioners in situations where they felt their actions violated values of personal autonomy.

> *So, when government started mandating people getting vaccines in order to keep their jobs, obviously I’m pro-vaccines but it was super difficult going into work every day. I’d literally have people crying in the seat saying I don’t want this, but I can’t feed my kids unless I get this and so that was an exhausting time, right? Because you’re telling people like technically, I need your informed consent to give you this vaccine and they’re like, well, I don’t have a choice and like I would never actually be able to have that conversation with them and have them be like, you know, yeah, this is the right decision […] People didn’t actually have the choice, and as a healthcare practitioner, we didn’t actually always get what felt like informed consent from these people, right? Like they literally just sit there and cry and be like, well, I don’t want it but do it. [P002, Nurse, 2.5 years in public health and healthcare]*.

Practitioners were further pressured to meet unrealistic client quotas in vaccine-hesitant communities.

> *One person every 5 minutes. That is not enough time to see them, to chart, to give the vaccine and to do a robust informed consent, especially in a very vaccine hesitant geographical area. It was insane. [P011, Nurse, 3 years in public health, 12 years in healthcare]*.

Workplace violence was frequently mentioned and caused participants to lose trust and faith in people, a defining symptom of moral injury [27].

> *How am I gonna mitigate or manage this situation, with what I felt was, you know, impending violence. And I’m driving into work and again, this is the community I live in. This is where you know I’ve worked for so long […] and then I just like kind of lost it, because I though like, like what… where am I working? That I like… just the act of calling the cops was really impactful for me. And I was like, it just… yeah, that was kind of the last straw. That it was like, where am I working? And just disappointment in I think my employer, but also in the community that we had got to that stage and I don’t think… no one was really acknowledging that we are, we are just people doing their jobs. […]I think that feeling of being taken advantage of or it’s like I’ve lost…like felt a bit used or something by friends, family, community, my employer, and I’ve like, lost some faith in people or something because of it, you know? [P014, Nurse, 20 years in public health, 24 years in healthcare]*.

Beyond pandemic experiences, participants described distress from witnessing drug use. They questioned their role in harm reduction, supporting client autonomy to consent to use drugs but questioned whether clients truly consented or lacked choices due to systemic inequality. Witnessing this without any ability to influence systemic changes caused moral injury.

> *Watching someone self-harm for 8 hours a day, or however frequency, like there is some sort of distress where it goes against the code, like basic ethical principles of beneficence and do no harm. Because you’re watching someone do harm, even though it’s their own informed consent that they’re doing. But then at a higher level, you’re questioning how much consent is it when they’re coming from a place where there’s social determinants of health, they may not have many options, or there’s no other place for them to find the respite from their very distressed life other than drugs. And like it’s a system failure, so that’s why they’re doing this. And then you’re witnessing it. [P009, Nurse, 17 years in public health, 24 years in healthcare]*.

Moral injury extended into personal lives, in some cases leading to anti-anxiety medication use, substance misuse and the need for counselling and cognitive behavioural therapy. Ultimately public health practitioners frequently chose to leave public health work as the only recourse to address moral injury. 80% of participants indicated that they had taken medical or sabbatical leave, planned on leaving their immediate job or had recently left a front-line position since the pandemic.

### Impacts of experience on moral distress

Experience, especially in public health, shaped participants’ approaches to work, advocacy, and sources of moral distress. Those with at least 10 years in healthcare and 5 in public health often accepted that not everything can be changed, valuing ‘small wins’ in individual’s lives as much as systemic changes. They used indirect advocacy methods, such as circumnavigating organizational structures or involving others from different fields when constrained. One participant described engaging retired teachers to oppose cuts affecting a key community partner in health education.

Moral outrage was present in all participants, but responses evolved with experience. Less experienced practitioners demonstrated ungrounded moral outrage, feeling anger without ability to channel it. Those with over 10 years’ experience emphasized specific advocacy methods, suggesting experience grounds moral outrage into positive action [32]. They spoke assertively against unjustified decisions, less deterred by reprimands. Those reflecting on early career moral distress noted their inexperience led them to accept structural constraints they would now challenge through constructive action.

Experience also fostered a commitment to mentoring. Experienced practitioners supported newer staff by speaking up to management, risking reprimands to reduce other’s moral distress.

> *I will always you know, if I have a colleague who’s like, can you bring it up at this meeting? OK, like you know, I’m not afraid to speak out, like, you know, people are. I’m not afraid to raise my hand and say my opinion. It’s probably gotten me in trouble multiple times, but I mean I do think […] it’s important to bring about change and to, you know, to reduce the moral distress when people feel like they’ve been heard. [P008, Nurse Practitioner, 11 years in public health, 22 years in healthcare]*.

### Inequity as a systemic cause of moral distress

Participants who disclosed a minority population identity through gender or race expressed notable differences in how they spoke about their work. Their affinity with minority communities led to feelings of increased responsibility, exacerbating experiences of moral distress.

> *And I was wishing for some other people to fight for me, and to have that activism view of their work and really being involved, but people were mostly just there to do their nine to five and go home and not really care that much because they were not really like people from the community. It didn’t matter to them the way it mattered to me because it was my community. It was my health. It was my people’s health and that mattered to me. So that was very hard and isolating in that experience. Having to fight from inside when I just want to fight the outside with the limited resources I already have. [P004, Social Worker, 5 years in public health and healthcare]*.

Participants selected specific work portfolios to support their communities but for them moral distress was compounded by discrimination and their interstitial social position as both witness and community member.

> *What drew me to do my masters and to choose my subject was rooted in the like conflict and moral distress I was living as a member of my marginalized community, working in Community settings where it’s supposed to be an empowering place, a place where you can do like grass work and really involve yourself with your community and work for the best. But I experienced a lot of heterosexism. I experienced a lot of structural oppression by CIs, white, straight people in positions of power not being allies but being oppressive. Not knowingly but being oppressive in the ways they manage the place. So, I think that was the first tension that caused me moral distress, cause I begin to be really confused between my identity as a queer person working in queer community settings with all the HIV and AIDS activism behind me, inspiring me and really pushing me to do that work. And the professional identity, professional expectations like hierarchical those types of forces. […] I really thought going into that work would be an empowering vessel, and I have these allies that was working with me and listening to what we wanted to do, but the truth was the contrary. I really liked performative active allyship, but when it matters the most, you’re not being supported. I’ve experienced transphobia by the same people. [P004, Social Worker, 5 years in public health and healthcare]*.

They also described career constraints due to racism and discrimination as morally distressing.

> *So on one hand, we’re here talking about health equity and we’re an equitable organization, but internally, people were being discriminated against, you seeing the layoffs happen and you question how come certain people are getting laid off, they claim, you know, like. […] And there was distress for these racialized staff because, and I think that’s a subset where you feel like you are wanting, you’re there with 100%, but you know the system, the oppression, completely excludes you from certain opportunities. [P006, Nurse, 2.5 years in public health, 10 years in healthcare]*.

These findings show how moral distress and injury interplay differentially within the public health workspace. Positioned at both individual and population levels, practitioners are simultaneously witnesses and participant in systems marked by inequity and discrimination. Valuing social justice and advocacy, public health practitioners attempt to address broad determinants of health such as poverty and marginalization [10,11], while also working directly with individuals and communities. This leads to moral distress as they are unable to improve these systems while witnessing impacts on clients and communities. Moral outrage develops from distress, culminating in moral injury when systemic inequities remain unaddressed.

## Discussion

Systemic powerlessness was a key driver of moral distress among public health practitioners in this study. Participants described witnessing systemic discrimination and felt a moral imperative to advocate for structural changes. Advocacy was a central theme across interviews and was seen as integral to the public health role. However, once within systems that espoused advocacy and justice, many practitioners found them constrained and even oppressive. Evidence-based policymaking was often sidelined, limiting practitioners’ ability to respond effectively to community needs.

The work environment and perceived value of public health impacted moral distress. Public health work environments extend beyond hospitals or institutions and include social, political and ideological contexts. While patient centred care is a core value of healthcare, little attention is given to the social and systemic conditions affecting health [10]. Chronic underfunding of prevention and promotion work, especially for vulnerable populations, left practitioners feeling undervalued and contributed to moral distress as they witnessed health inequalities without ability to improve them.

Moral distress was heightened for practitioners serving their own communities, experiencing distress both as providers and community members affected by systemic inequities.

Moral injury was common, with distinct exacerbation during the pandemic. While pre-pandemic morally injurious events were discussed, most moral injury was framed around the pandemic. Inability to drive change in a politically hostile and undervalued public health context led to symptoms of moral injury such as a loss of trust in humanity.

With experience, some participants turned their moral distress into positive action overtime, as they channelled their anger from moral outrage into advocacy. Still, this did not mitigate the personal toll. Many eventually left public health or frontline roles removing them from the downstream witness advocating for upstream changes, which was the core of moral distress experiences explored in this study.

Moral distress is well-documented in clinical contexts but underexplored in public health[8].^8^ The pandemic increased focus on public health, revealing moral distress from political dominance over science [6–8,33]. Some research identified increasing moral distress for Canadian community nurses witnessing worsening health disparities [7]. Practitioners feel moral distress when promoting guidance they believe to be inadequate, or when system policies restrict patient care [7,8,33,34].

Evidence links moral distress to unethical work environments [32,33,35]. The culture of the workplace, including feelings of being valued, has a huge impact on employees and their moral distress [8].

While many studies on moral distress collect gender and race data, few analyze it meaningfully [36]. Those that do highlight racisms, class and power relationships, ethnic affiliation and cultural beliefs as key contributors [36].

Most moral injury research originate from military settings [27,37,38]. Public health remains underexamined, with existing studies emphasizing quantification over deeper understanding. Some evidence links workplace violence to moral injury [6,22,39].

This study confirms that systemic powerlessness, political interference, work environments including communication and hierarchical structures, and the devaluation of expertise contribute to moral distress in public health. It further confirms that the pandemic exacerbated existing moral distress in public health practitioners, triggering moral injury in many. What distinguishes this study is the in-depth interrogation into these experiences, with analysis of intersectionality, considering years of experience, gender and ethnicity and how these contributed differentially to moral distress. Participants’ varied backgrounds and years in the field provided insights into how experience affected advocacy, coping and peer support.

Importantly, this study contrasts pre and post pandemic moral distress experiences and allows us to do so explicitly in a Canadian context. It highlights that the pandemic amplified existing, unresolved moral distress, and potentially pushed distress into injury for many. It contributes firsthand accounts of moral distress experiences, and the methods developed over time to address it. This allows us to consider potential strategies for mitigating long-term harm. While those with more experience accepted informal peer support roles to reduce moral distress for newer colleagues, establishing formal peer networks or creating anonymous or non-punitive pathways to provide concerns to leadership could be explored for their impact on moral distress. Further focus on communication from front-line non-leadership staff to leadership and vice versa should occur in public health. Ensuring the voices of those with experience are listened to and acted on and ensuring essential information that could impact patient care is delivered to those who need to utilize it should be done in systematic and consistent ways, including during pandemic and crisis times. Ensuring racism and any form of discrimination are addressed and rooted out across any public health organizations will help reduce the double burden those from marginalized communities identified while facing discrimination both from within and outside an organization. Supporting new staff in public health to learn about advocacy channels and methods, either through trainings, educational sessions or connections with professional organizations who support advocacy could encourage more positive action from moral outrage.

We extend the concept of organizational hierarchy as a trigger for moral distress beyond clinical contexts into public health work [8]. Hierarchy operates across health systems and political structures, not solely within institutional boundaries. The hierarchical structures that enhance systemic powerlessness and the active silencing, defunding and devaluation of expertise that public health practitioners faced when working within politically challenging environments, both before and during the pandemic, is indicative of the extent of inequality and discrimination they are witness to. Witnessing resource allocation follow systemic fault lines of racism and discrimination without being able to affect change, both during and outside the pandemic, was especially morally distressing.

It is important to note that some of the topics discussed in this study were of a sensitive nature. Participants had to be re-assured of the confidentiality of their conversations, and some expressed fear of being identified, asking for certain quotes or stories to be excluded. This is in itself an interesting finding and highlights the extent that intimidation, fear, oppression, and undervaluing leads to feelings of powerlessness in public health structures.

This study contributes new evidence of moral injury in public health and its role in prompting career exits. The pandemic was a breaking point, but moral distress had long preceded it.

### Strengths and limitations

While the sample size was small, this study achieved saturation, capturing core themes of moral distress among Canadian public health professionals. While not generalizable, findings align with trends in other high-income settings.

As participants self-identified, we cannot determine how widespread moral distress is, nor how it differs quantitatively across public health sectors. However, participants worked across a range of public health sectors and reported consistent themes. Despite efforts to recruit ethnically diverse populations, few beyond white and female self-identified. Reasons are unknown but may include fatigue, disillusionment, or systemic distrust – especially among marginalized populations.

Data were collected during three months in 2023, shortly after the World Health Organization declared COVID-19 over as a global health emergency [40]. While timing shaped reflections, interviews included pre- and post-pandemic experiences, enabling comparison.

## Conclusion

Moral distress is present across healthcare, but in public health it is shaped by the field’s dual focus on upstream and downstream work, and emphasis on advocacy and justice [10,11]. This study shows that although the pandemic intensified moral distress, it did not originate it. Drawing on practitioners’ own coping strategies may inform long-term solutions. Valuing expertise, reducing political interference, and fostering safe work environments may help mitigate moral distress and prevent moral injury in public health.

## Data Availability

The data underlying this article cannot be shared publicly for the privacy of the individuals that participated in the study. The data will be shared on reasonable request to the corresponding author.

## Acknowledgements

We thank Dr. Sarah Smith [London School of Hygiene and Tropical Medicine], the study participants, THE BRIDGE staff, and the Canadian Public Health Association.

## Supporting information

S1 File. Supplementary File Discussion Guide. S1_File.docx

